# Development and validation of a rapid, single-step reverse transcriptase loop-mediated isothermal amplification (RT-LAMP) system potentially to be used for reliable and high-throughput screening of COVID-19

**DOI:** 10.1101/2020.03.15.20036376

**Authors:** Minghua Jiang, Weihua Pan, Amir Aratehfar, Wenjie Fang, Liyan Ling, Hua Fang, Farnaz Daneshnia, Jian Yu, Wanqing Liao, Hao Pei, Xiaojing Li, Cornelia Lass-Flörl

**Affiliations:** Department of Laboratory Medicine, The Second Affiliated Hospital and Yuying Children’s Hospital of Wenzhou Medical University, Wenzhou, China; Shanghai Key Laboratory of Molecular Medical Mycology, Shanghai Institute of Mycology, Shanghai Changzheng Hospital, Second Military Medical University, Shanghai, China; Westerdijk Fungal Biodiversity Institute, Utrecht, The Netherlands; Department of Laboratory Medicine, Pinghu Second People’s Hospital, Zhejiang, China; Department of Laboratory Medicine, Pudong New Area People’s Hospital, Shanghai, China; Department of Laboratory Medicine, Wuxi No. 5 People’s Hospital, Jiangsu, China; Department of Dermatology, Affiliated Hospital of Hebei University of Engineering, Hebei, China; Institute of Hygiene and Medical Microbiology, Medical University of Innsbruck, Innsbruck, Austria

**Author notes:** **Correspondence:** Xiaojing Li, Hao Pei. Minghua Zhu, Weihua Pan and Amir Aratehfar equally contributed to this study.

**Keywords:** RT-LAMP, COVID-19, qRT-PCR, SARS COV-2

## Abstract

**Objectives:** Development and validation of a single-step and accurate reverse transcriptase loop-mediated isothermal amplification technique (RT-LAMP) for rapid identification of SARS COV-2 relative to commercial quantitative reverse transcriptase real-time PCR (qRT-PCR) assays to allow prompt initiation of proper medical care and containment of virus spread.

**Methods:** Primers showing optimal *in-silico* features were subjected to analytical sensitivity and specificity to assess the limit of detection (LOD) and cross-reaction with closely- and distantly-related viral species, and clinically prominent bacterial and fungal species. In order to evaluate the clinical utility, our RT-LAMP was subjected to a large number of clinical samples, including 213 negative and 47 positive patients, relative to two commercial quantitative RT-PCR assays.

**Results:** The analytical specificity and sensitivity of our assay was 100% and 500 copies/ml when serial dilution performed in both water and sputum. Subjecting our RT-LAMP assay to clinical samples showed a high degree of specificity (99.5%), sensitivity (91.4%), positive predictive value (97.7%), and negative predictive value (98.1%) when used relative to qRT-PCR. Our RT-LAMP assay was two times faster than qRT-PCR and is storable at room temperature. A suspected case that later became positive tested positive using both our RT-LAMP and the two qRT-PCR assays, which shows the capability of our assay for screening purposes.

**Conclusions:** We present a rapid RT-LAMP assay that could extend the capacity of laboratories to process two times more clinical samples relative to qRT-PCR and potentially could be used for high-throughput screening purposes when demand is increasing at critical situations.

## 1 Introduction

A new virus causing pneumonia-like infection, COVID-19, which was found in Wuhan, Hubei Province, China, and to be linked to a seafood market has caused a serious crisis worldwide (1). Almost two months after the first report, COVID-19 severe outbreaks were reported in numerous countries and became a public health priority in the world (World Health Organization, Situation Report 48). As of March 17, 2020, COVID-19 cases are found in 150 countries/regions and infected 167,515 patients, among whom 6,606 died (World Health Organization, Situation Report 56). The latest phylogenetic analysis studies designated the etiologic agent of COVID-19, SARS COV-2 (2).

The virulent nature of this virus and its high rate of transmissibility warrants robust, rapid, sensitive, specific, and quantitative diagnostic tools to supplement clinical symptoms aiding clinicians to confidently rule in and rule out patients. Moreover, such a diagnostic tool will help with preventing spread of virus by identifying the infected cases and can monitor the health status of infected patients by quantifying the viral load. Center for Disease Control was the first to develop a quantitative reverse transcriptase real-time PCR (RT-PCR), which later became the gold standard technique (https://www.cdc.gov/coronavirus/2019-ncov/about/testing.html). Subsequently, a Chinese group used a RNA-based metagenomics next generation sequencing (mNGS) to diagnose the viral RNA from the clinical samples of two patients (3). However, the requirement for advanced technology and skilled personnel and long turn-around time (24 hours) are not feasible for local and referral laboratories. Therefore, a colorimetric loop mediated isothermal amplification, also known as LAMP, was developed to obviate the need for expensive technologies, e.g. real-time PCR and NGS, as well as to shorten the turn-around time to up to 40 minutes (4). However, this assay was a qualitative one and also only the swab samples from limited number of patients (*n*=7) were included for testing (4). Most recently a newer generation of single step RT-LAMP were developed to detect SARS COV-2, but these assays were not challenged with real clinical samples obtained from COVID-19 positive patients (5, 6). Therefore, we developed a sensitive, specific, and rapid RT-LAMP assay and its performance was challenged by an extensive number of confirmed COVID-19 (*n*=47) and negative patients (*n*=213) relative to qRT-PCR assays approved by two Chinese Food and Drug Administration (qRT-PCR NMPA). Altogether, we present a rapid and reliable diagnostic tool that potentially could be deployed for high-throughput screening applications in referral and local laboratories.

## 2 Materials and methods

### Target selection

According to *Guidelines for prevention and control of Covid-19 (Fourth Edition)* issued by National Health Commission of the PRC on 2020.2.26, open reading frame 1ab (ORF1ab) or nucleocapsid protein (N) were recommended for designing diagnostic assays detecting SARS-HCoV-2 from clinical samples. Therefore, ORF1ab and N sequences of SARS-Cov-2, its close related coronavirus species (HCoV-NL63, HCoV-OC43, HCoV-229E and HCoV-HKU1), and other viral species, namely Adenovirus, Respiratory syncytial virus A, Human parainfluenza 2 virus, Human parainfluenza 3 virus, H1N1 influenza virus, H5N1 influenza virus, H7N9 influenza virus, H9N2 influenza virus, Mycoplasma pneumoniae and Influenza B virus, were downloaded from GenBank (https://www.ncbi.nlm.nih.gov/genbank/) to select the most specific target region. Genenious v11.1.14 was used for alignment analysis and to find the most specific region for designing LAMP primers. LAMP Designer (PREMIER Biosoft International, San Francisco, CA) was used for primer design. Designed primers were subjected to BLAST (https://blast.ncbi.nlm.nih.gov/Blast.cgi) and the specific candidates were used for analytical sensitivity and specificity testing (Table 1). Primers were synthesized by Sangon Biotech Co., Ltd. (Shanghai, China).

**Table 1.**
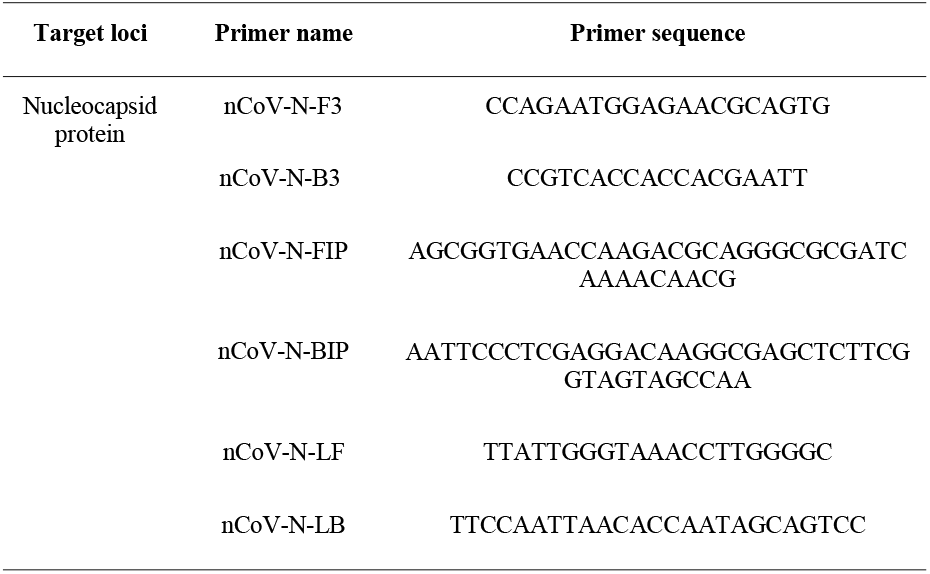
Primers and probes successfully detected SARS COV-2.

### Analytical sensitivity and specificity testing

Analytical sensitivity and specificity testing was performed in a P2 lab and in order to mimic the real viral particles we purchased pseudotyped SARS-CoV-2 assay system containing ORF1ab part sequence, N gene and E gene (DAAN gene Co. Ltd, Guangzhou, China). RNA of pseudotyped virus were extracted using EZ-10 Spin Column Viral Total RNA Extraction Kit (Sangon Biotech Co.,Ltd. Shanghai, China). Serial dilutions with the magnitude of log10 containing 50*10^6^ cell/ml to 50*10^0^ cell/ml pseudotyped virus were performed to determine the limit of detection (LOD). Serial dilution testing was performed in both RNase/DNase free molecular grade water and sputum sample collected from a COVID-19 negative healthy individual. Reproducibility of our LAMP assay (linearity=R^2^ value) was assessed by separate serial dilution testing on three occasions, each performed in duplicate. Signal intensity and the time to obtain decent amplification curves were recorded and R^2^ values ≥0.98 were considered reliable amplification.

Specificity testing included nucleic acid of HCoV-OC43, HCoV-HKU1, HCoV-229E, HCoV-NL63, Adenovirus, Respiratory syncytial virus A, Human parainfluenza 2 virus, Human parainfluenza 3 virus, H1N1 influenza virus, H5N1 influenza virus, H7N9 influenza virus, H9N2 influenza virus, Mycoplasma pneumoniae, Influenza B virus (Bdsbiotech Co. Ltd, Guangzhou, China). Moreover, HeLa cells (TechStar Co. Ltd, Jiangsu, China) and genomic DNA of clinically prominent bacteria or fungal species, including *Staphylococcus aureus, Mycobacterium tuberculosis, Legionella pneumophila, Candida albicans, Candida glabrata, Candida tropicalis, Aspergillus fumigatus, Cryptococcus neoformans* were used for specificity testing (provided by Shanghai Institute of Medical Mycology, Shanghai Changzheng hospital). LAMP incubation time was set to 60 minutes to observe both limit of detection and cross-reactivity (LAMP conditions are mentioned in clinical evaluation section). The reaction endpoint time was set in a way to detect the lowest possible copy number of virus without any cross-reaction.

### Evaluating LAMP assay tolerance against wide range of inhibitors

Clinical samples obtained from patients contain a wide range of inhibitors impairing the efficacy of diagnostic assay. Therefore, the tolerance of our LAMP assay was assessed when 500 copy/ml of simulated viral particles were mixed with human blood, mucin, β-adrenergic bronchodilator, Tamiflu, dexamethasone, adrenaline.

### Clinical validation

Clinical validation engaged two clinical centers, namely The Second Affiliated Hospital and Yuying Children’s Hospital of Wenzhou Medical University, Wenzhou, China (Center one), and the Wuxi Infectious Diseases Hospital, Wuxi, China (Center two). Each center used a different qRT-NMPA assay as a gold standard technique. SARS-CoV-2 kit from Shanghai BioGerm Medical Biotechnology Co. Ltd, (NMPA approval number 20203400065, with LOD of 1000 copies/ml, Ct cut-off 38), and a kit from DAAN Gene Co., Ltd (NMPA approval number 20203400063, with LOD of 500 copies/ml, cut off Ct value of 40) were used in center one and center two, respectively. Positive patients were divided into two groups by physicians, namely suspected and confirmed. Those suspected were isolated and all became positive. The ethics committees of both centers approved the study. Emergency patients (outpatients) with fever of unknown origin or inpatients diagnosed as COVID-19 or other diseases were enrolled and samples such as sputum, swabs and tears were used for evaluation. ABI 7500 were used for amplification and data analysis in both centers.

The final LAMP reaction was 25 µl and contained 21.9 µl buffer solution (20 mM Tris-HCl pH 8.8, 10 mM (NH4)2SO4, 120 mM KCl, 2 mM MgSO4, 0.1% Tween 20), 8 U Bst DNA polymerase (New England Biolabs (Beijing) ltd, Beijing, China), 0.5 U AMV Reverse Transcriptase (Takara Bio Inc, Dalian, China), 2 µl RNA template, 1.6 μMFIP/BIP primers, 0.2 μM F3/B3 primers, 0.4 μM LF/LB primers, 7 mM MgSO4 (Sangon Biotech Co., Ltd., Shanghai, China), 0.8M betaine (Sangon Biotech Co., Ltd., Shanghai, China), 1.4 mM each dNTP (Takara Bio Inc, Dalian, China), 0.5 μM SYTO-9 (Invitrogen Trading, Shanghai) Co., Ltd, Shanghai, China). LAMP reactions were incubated at 63^º^C for 30 mins in ABI 7500 machine and florescent data were collected each minute. RT-PCR and RT-LAMP tested separately by two assessors, final results were recorded and were compared with one another.

## 3 Results and discussion

The whole workflow of our study from *in-silico* analysis to analytical evaluation and clinical validation is depicted in Figure 1. Nine and six LAMP primer systems were designed and evaluated *in-silico*, but only the six primers showed the highest sensitivity and specificity, which used in the next steps (Table 1). Primarily, our assay was meant to be quantitative and it showed an optimal reproducibility when tested in analytical evaluation step using armored viral particle diluted in water (R^2^ value □0.99) and sputum sample (R^2^ value □0.83). Analytical sensitivity yielded reliable LOD of 500 copies/ml less than 30 minutes regardless of matrix used for serial dilution (Figure 1). Of note, our assay could detect 50 copies/ml, but some replicates showed unstable amplification. Therefore, we considered the LOD of 500 copies/ml. Analytical specificity was 100% when used a wide range of closely- and distantly-related viral species, prominent fungal and bacterial species, and human DNA. Moreover, analytical evaluation included a wide range of inhibitors and 500 copies of the simulated viral particles were successfully detected below 30 minutes (Figure 1). In order to evaluate the performance of our assay in clinic, we provided our assay and respective instructions to two clinical centers (Figures 1, 2). In total, 168 patients from center 1, among which 35 confirmed COVID-19 cases, and 92 patients from center 2, among which 12 patients were confirmed COVID-19 cases, were recruited. One asymptomatic patient tested positive by qRT-PCR (Ct values 37) and by our RT-LAMP was categorized suspected by in-charge physician and few days later became positive. Four patients tested positive by qRT-PCR were negative by our RT-LAMP and one patient tested negative by qRT-PCR was positive by our assay (Figure 1 and Supplementary Table 1). Subsequently, our RT-LAMP assay showed the sensitivity, specificity, negative predictive value, and positive predictive value of 91.4%, 99.5%, 98.1%, and 97.7%, respectively (Supplementary Table 2 and 3). The fact that our assay could not detect four positive patients was owing to using 2.5 less RNA input (2 µl) relative to qRT-PCR (5 µl). In the future, we will try to use various RNA input volume (5, 8, and 10 µl) to observe if we could obtain a higher sensitivity. Although our RT-LAMP assay was developed to be quantitative, we could not find any pattern and association between the time to positivity by our RT-LAMP assay and the Ct values reported by qRT-PCR when using clinical samples. Therefore, we considered our assay a qualitative one. This fact will show that the analytical valuation should be always accompanied by clinical validation to observe the real capabilities of a given assay and that the results obtained in analytical evaluation step are not always reflected in real-life.

**Figure 1.**
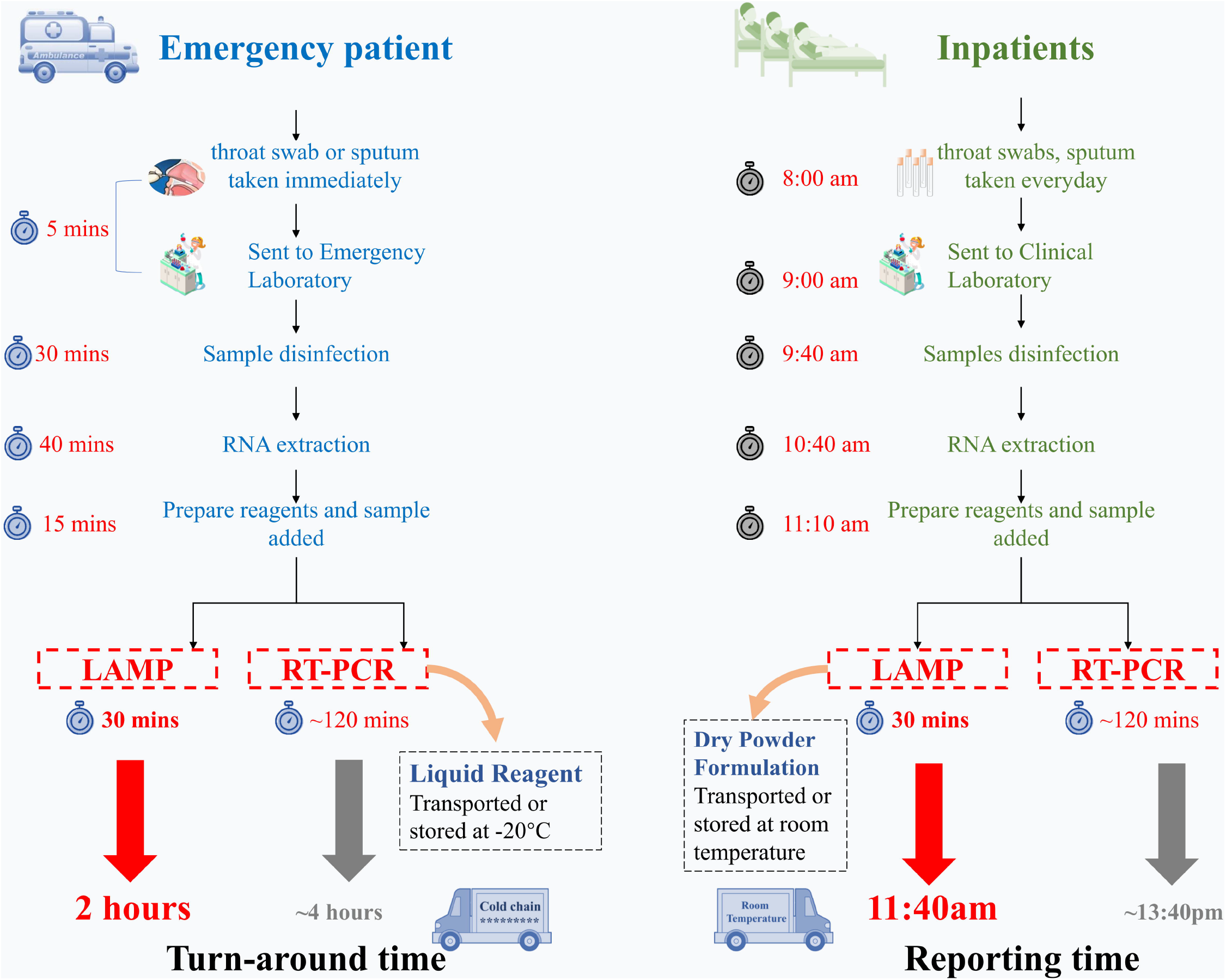
Our assay was comprehensively evaluated at three steps, including in-silico analysis, in-vitro analytical analysis, and clinical validation.

**Figure 2.**
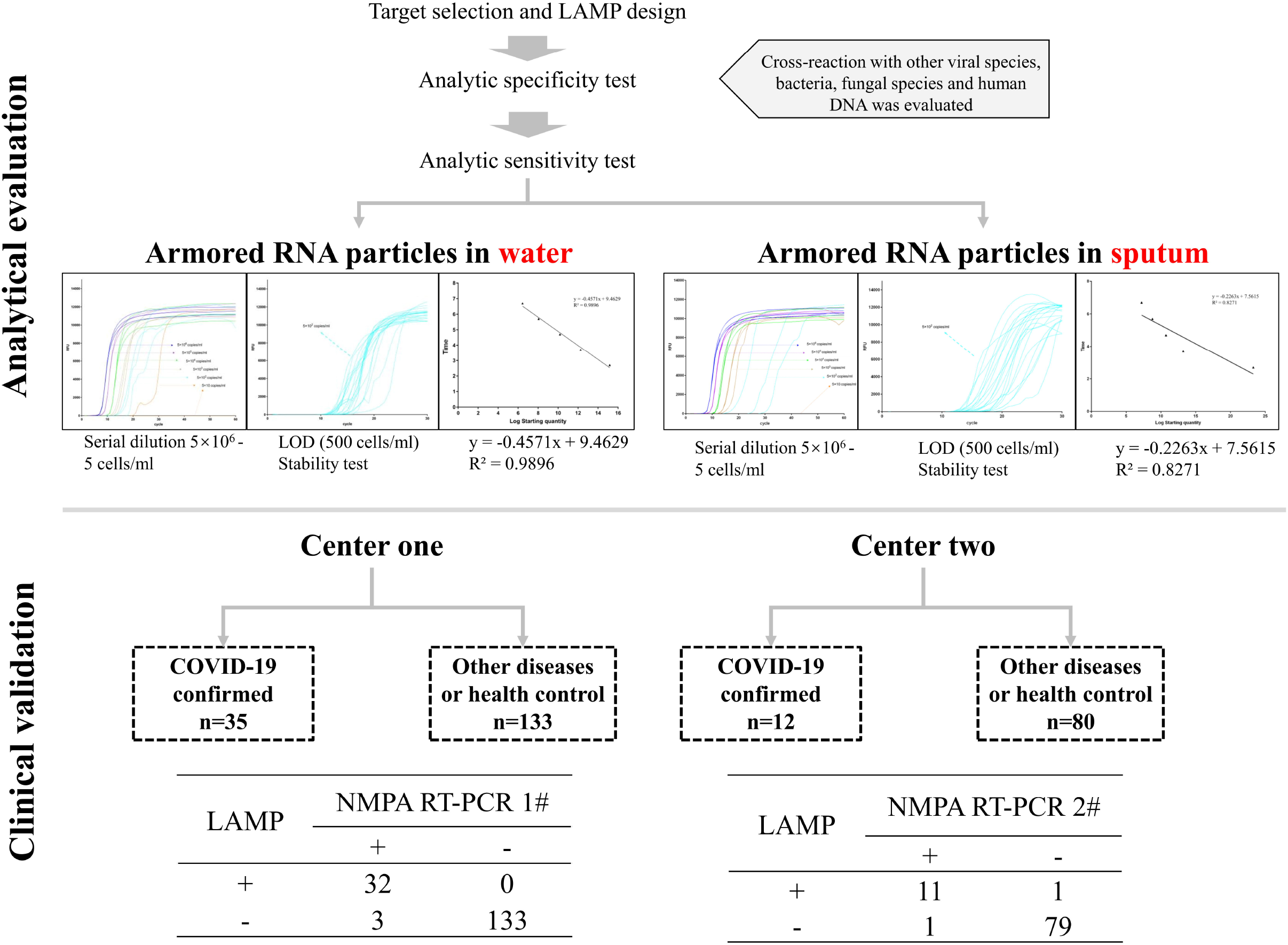
Workflow comparison of our RT-LAMP assay relative to qRT-PCR for emergency cases (outpatients) and inpatients. Our RT-LAMP assay is 2 to 2.5 times faster than the qRT-PCR assays and can be shipped at room temperature.

Our assay has several advantageous compared to qRT-PCR. First, our RT-LAMP assay is two times faster relative to qRT-PCR (Figure 2) and given the optimal diagnostic features could be used as a reliable screening method in local and referral laboratory to keep up with the increasing demand of suspected patients in critical situations. Secondly, our assay does not need the clod chain and could be shipped at room temperature (Figure 2).

In conclusion, we present a rapid RT-LAMP assay that allows processing 2 to 2.5 more clinical samples relative to CDC RT-PCR, which is indicative of its capacity to be deployed for high-throughput screening applications in local and referral laboratories.

We admit that our assay does not has the quantitative aspect of qRT-PCR and its sensitivity requires improvement. These two limitations will be the subject of future investigation. Moreover, we will try to use simple and fast nucleic acid extraction procedures (7) that only uses heat that will further decrease the turn-around-time.

## Data Availability

The data used to support the findings of this study are available from the corresponding author upon request.

## Ethics Statement

The protocol was approved by the Ethics and Research Committee of The Second Affiliated Hospital and Yuying Children’s Hospital of Wenzhou Medical University, Wenzhou, China, and the Wuxi Infectious Diseases Hospital, Wuxi, China

## Author Contributions

JMH, PWH and AA participated in primer design, LAMP optimization, data collection, and drafted the manuscript. LXJ and PH participated in designing this study and revising the manuscript. WJF, LLY, FH participated in collecting clinical samples. All authors contributed to the writing of the final manuscript.

## Supplementary Material

**Appendix Table 1**. Signal intensity and time corresponded to each serial dilution obtained during three separate experiments. Each experiment was performed in duplicate.

**Appendix Table 2**. False positive and false negative results reported by our RT-LAMP assay, when compared to qRT-PCR.

**Appendix Table 3**. Diagnostic feature of our RT-LAMP assay when compared to qRT-PCR.

## Reference

1. Ma X, Ph D, Wang D, Ph D, Xu W, Wu G, Gao GF, Phil D, Tan W, Ph D. 2020. A Novel Coronavirus from Patients with Pneumonia in China, 2019. New England Journal of Medicine 727–733.

2. Wu Y, Ho W, Huang Y, Jin D, Li S, Liu S, Liu X, Qiu J, Sang Y, Wang Q, Yuen K, Zheng Z. 2020. SARS-CoV-2 is an appropriate name for the new coronavirus. The Lancet 6736:2–3.

3. Chen L, Liu W, Zhang Q, Xu K, Ye G, Wu W, Sun Z, Liu F, Wu K, Zhong B, Mei Y, Zhang W, Chen Y. 2020. RNA based mNGS approach identifies a novel human coronavirus from two individual pneumonia cases in 2019 Wuhan outbreak. Emerging microbes & infections 1751.

4. Zhang Y, Odiwuor N, Xiong J, Sun L, Nyaruaba RO, Wei H, Tanner NA, Tanner N. 2020. Rapid Molecular Detection of SARS-CoV-2 (COVID-19) Virus RNA 2.

5. Park G, Ku K, Beak S, Kim SJ, Kim S Il, Kim B. 2020. Development of Reverse Transcription Loop-mediated Isothermal Amplification (RT-LAMP) Assays Targeting SARS-CoV-2.

6. Lamb LE, Bartolone SN, Ward E, Chancellor MB. 2020. Rapid Detection of Novel Coronavirus (COVID-19) by Reverse Transcription-Loop-Mediated Isothermal Amplification.

7. Myhrvold C, Freije CA, Gootenberg JS, Abudayyeh OO, Metsky HC, Durbin AF, Kellner MJ, Tan AL, Paul LM, Parham LA, Garcia KF, Barnes KG, Chak B, Mondini A, Nogueira ML, Isern S, Michael SF, Lorenzana I, Yozwiak NL, MacInnis BL, Bosch I, Gehrke L, Zhang F, Sabeti PC. 2018. Field-deployable viral diagnostics using CRISPR-Cas13. Science 360:444–448.

